# Factors Associated with Career Ambitions in General Internal Medicine: Insights Into Gender Disparities in Leadership Aspirations

**DOI:** 10.1101/2025.05.18.25327847

**Authors:** Jeanne Moor, Lena Woodtli, Christine Baumgartner, Karolina Kublickiene, Sven Streit, Christa Nater

**Author notes:** Correspondence to: Jeanne Moor, MD, Institute of Primary Health Care (BIHAM), University of Bern, Mittelstrasse 43, 3012 Bern, Switzerland.

## Abstract

**Background:** The existence of a “leaky pipeline” in leadership for medicine in Switzerland is apparent: 60% of residents and 51% of attending physicians are women; yet women make up only 32% of senior physicians and 16% of chief physicians. Here, we examined sex-specific and gender-sensitive factors affecting career ambitions for physicians in General Internal Medicine (GIM).

**Methods:** Using a cross-sectional survey among GIM physicians in Switzerland, we assessed women’s and men’s career ambitions; workplace-related factors including professional network, perceived supervisor’s support, workplace inclusiveness, and gender-related discrimination; personal factors such as Physician Well-being index and family situation; and demographics. The outcome was the probability of aspiring to a senior position (i.e., senior or chief physician) at a hospital. Chi-squared tests and multiple logistic regression identified the factors associated with this career aspiration and sex-disaggregated analysis tested for sex differences.

**Results:** The sample included 624 physicians, of which 40% were men (mean age = 37 years, standard deviation [SD] = 11 years), and 60% women (mean age = 35 years, SD = 10 years). Their workplace was mostly in hospitals (76%) or in private practice (21%). During medical school, the career aspiration of men and women did not differ (men vs. women: 14% vs. 12%, p=0.62). Yet, in their current aspiration towards leadership, men more frequently reported high aspirations than women (17% vs. 11%) in univariable analysis (p=0.03) and odds ratio [OR]: 1.74, 95% confidence interval [CI]; 1.06-2.85, after multivariable adjustment. Higher career aspiration was present among those who were more satisfied with their work (OR: 3.26, 95% CI: 1.56-7.73). In sex-stratified analyses, men with career aspiration were less likely to have a pathological Physician Well-being index than men without this aspiration (OR: 0.12, 95% CI: 0.02-0.50). Women with career aspiration were more likely to report gender discrimination, or to face a negative view on parenthood by their supervisors, compared to women without career aspiration (OR: 7.48, 95% CI: 1.71-39.0). However, having children or having adequate childcare were not associated with career aspiration among respondents of any sex.

**Conclusion:** Both female and male physicians had similar career aspirations at the beginning of their career during medical school, while the sex difference emerged as their careers progressed and different challenges emerged. Sex-specific associations among the women, with supervisors’ unfavorable views on parenthood and gender discrimination relating to leadership aspirations, provide evidence-based knowledge for workplace-tailored strategies to close the leak in the pipeline towards leadership positions in medicine.

## Introduction

Women have become the majorities of medical students and residents in many European countries in the last decades.(1–3) However, their advancement for leadership positions is still far from equal. Women remain underrepresented among leadership in medicine both in the U.S. (4) and in most European countries.(1,5,6) In fact, women make up fewer than 20% of department chairs and deans.(6– 9) This constitute a gender gap in leadership linked to a “leaky pipeline”, a term coined over 20 years ago to describe the attrition of women and minority groups from a career in science after having received a bachelor’s degree.(10,11) Since then, “leaky pipeline” has mostly been used to describe the progressively decreasing fraction of female academics with advancing hierarchical position, especially so in medicine.

Known challenges for female physicians and physician-mothers include domestic and/or childcare responsibilities, work-life balance, lack of adequate mentors and role models, receiving fewer career support, and discrimination or sexual harassment due to gender and pregnancy.(12–14) Moreover, the relationship between an unsupportive workplace environment and declining career ambitions has been observed in non-medical professionals.(15) In Switzerland, scientists considered the lack of compatibility between family and work life as a key reason not to pursue an academic career.(16) Yet, it remains an open question how personal and workplace-related factors may affect career ambitions for physicians of the current General Internal Medicine (GIM) workforce.

Leadership aspirations and the representation of women in leadership positions in medical careers warrant further research due to existing knowledge gaps about the importance and relevance of personal and workplace-related factors that could better serve to identify and understand the priorities and/or obstacles of future health care providers. Some factors could be modifiable and help to improve the participation of women in leading positions in medicine. Therefore, we aimed to describe and compare past and current career aspirations of the Swiss GIM workforce in both women and men. Secondly, we determined potential factors associated with high career aspirations in the Swiss GIM workforce as a whole and specifically in women.

## Methods

### Design, setting and participants

This study reports on a cross-sectional web-based survey that was administered to GIM physicians in Switzerland. The participants were contacted via the head or administration of GIM departments in 14 Swiss hospitals, 6 ambulatories or primary care institutes, and via newsletter announcements by the Swiss Society of General Internal Medicine and the Swiss Young General Practitioners Association (JHaS), as well as by an advertisement in the Swiss journal titled “Primary Hospital Care”.

The study was waived and exempted from full review by the ethics committee of the canton of Bern under the identifier: Req-2021-01085. All participants were informed about the purpose of this study and gave informed consent for anonymous participation in the survey.

### Data collection

The survey was available from December 2021 to April 2022 and covered the following topics:

Outcomes of the study were: Aiming for a leading position, i.e. the aim of becoming a senior physician and/or chief physician, was the primary outcome of interest, and was assessed with a dichotomous item (Yes vs. No). As a secondary outcome, we chose aiming for an academic career. Those already holding a higher academic position or degree (professorship, lectureship or PhD) were excluded from the study population, and from the remainder all answering the question if they aim for an academic career with “yes” or “no” were analyzed. A third outcome was career ambition based on an abbreviated version of the 24-item Career Aspiration Scale-Revised (CAS-R).(17) The CAS-R is a 24-item questionnaire and has been validated in different populations.(17,28) Six questions were selected to assess career ambition, including two questions for each of the CAS-R domains chosen for their relevance for the medical profession. Each question was answered by a Likert scale ranging from 1 to 5 points, and the sum of responses of the abbreviated CAS-R questions were combined to a numerical career ambition score (CAS, minimum 6, maximum 30 points).

Further variables were collected as exposures of interest or potential confounders:

1. General demographic with information on participants’ age, Swiss language region, type of workplace and current job position, quota of employment, and relationship status;
2. Mental health assessment using the seven-item physician Well-being index (PWBI) to screen medical students for low mental well-being, burnout and fatigue.(18,19) The WBI was validated in large studies among U.S. physicians in the Mayo Clinic Program on Physician Well-being.(20,21) The standard cut-off value of ≥5 out of 7 PWBI points was considered as surrogate of impaired mental health.(20,21)
3. Job satisfaction was assessed by a single-item Likert question («How satisfied are you with your job?»), with responses ranging from 1 (dissatisfied) to 5 (satisfied).
4. Workplace culture assessment included four questions of the 19-items questionnaire by Hall et al. for measures of organizational culture.(22) The four close-ended questions assessing gender-related and general corporate inclusiveness. In addition, a self-developed question was added to determine if parenthood is viewed negatively at the workplace.
5. Family-related topics were identified and assessed using close-ended questions or Likert scale including having delayed having children, wishing or having children, age at birth of first child, the number of children, having adequate childcare, childcare duties and household duties compared with partner.(12)
6. Factors influencing career ambitions mentioned in the literature, including having a good network, having good mentoring and supervisor supporting an individual’s career (14).
7. Gender discrimination was assessed using 8 self-developed questions because of lack of suitable assessment instruments for physicians on this topic. The questions were partly inspired from a dedicated web-based tool and published literature.(23–26).

A pilot version of the questionnaire was tested in ten physicians for comprehensiveness and feasibility. The final questionnaire underwent back-and-forth translation from English to German or French and back by native speakers to ensure validity.(27) The survey was then offered to participants in German and French. The English version of the survey questions analyzed is available in Supplemental Table 1.

### Statistical analyses

Characteristics of study participants were presented as mean (standard deviation) or median (interquartile range) for continuous variables depending on their distribution, and n (%) for categorical data as n (%), with or without stratification by sex. Participants identifying as nonbinary were excluded from sex-stratified analyses because we considered them at risk for identification. Continuous variables were compared using Student’s t-tests or Wilcoxon rank sum test, as appropriate, and categorical variables using chi-squared tests. Next, we built univariable logistic regression models of factors associated with the primary outcome. Exposure variables were chosen based on the literature and included several personal and workplace-related characteristics.(14,29,30) For multivariable logistic regression, separate models were built for each exposure of interest, with Age, type of workplace and Swiss language region as a priori covariates and stratification by sex. For the second outcome, similar univariable and multivariable logistic models were established for aiming for an academic career. For the third outcome, univariable and multivariable linear regression models were constructed in a similar manner with CAS as the dependent variable. A subgroup analysis was performed for the main analysis of career aspirations in all parents with sex stratification. Finally, a sensitivity analysis was performed excluding general practitioners, because our definition of “leading position” in a hospital may deviate from career goals of general practitioners with high career ambitions. The obtained sample sizes for multivariable models were considered as suitable by an estimation of subjects per variable (SPV), with the literature referring to a minimum of 5 to 20 SPV or a fixed population size of at least 200 for multivariable modelling (31). This approach would deem a sample of 240 participants sufficient for inclusion of up to 12 covariates in regression models. All analyses were performed with R v. 4.1.3 and RStudio v. 2023.06.1. P-values <0.05 were considered as statistically significant. No adjustments were made for multiplicity.

## Results

### Demographic characteristics

The survey was completed by 684 physician respondents. For the present analysis, n=58 (8.5%) respondents currently holding a position as senior or chief physician were excluded, and n=2 (0.3%) respondents identifying as nonbinary were excluded from sex-stratified analyses. The characteristics of the analyzed sample of 624 physicians is displayed in Table 1, containing 237 (40%) men with a mean age of 37years (standard deviation [SD]11 years), and 387 (60%) women with a mean age of 35 years (SD 10 years). Work-place of 76% respondents was in hospitals and of 21% respondents in private practice. Current median work quota was 100% (IQR [interquartile range] 100, 100) in men and 100% (IQR: 80, 100) in women (p<0.001). The majority of respondents were in a relationship (88% of men and 80% of women, p<0.01). Men were less likely planning to delay (27% of men vs. 38% of women) or to have delayed having children than women (12% of men vs. 19% of women, p=0.002). Among workplace-related factors, gender discrimination questions were more frequently answered “yes” by women than men, but overall workplace inclusiveness was comparable between male and female respondents (Table 1).

**Table 1.** Physician characteristics.

| **Table 1. Physician characteristics** |  |  |  |  |
| --- | --- | --- | --- | --- |
|  | **N** | **Men, N=237** | **Women, N=387** | **p-value** |
| **Age (years), mean (SD)** | 624 | 37 (11) | 35 (10) | 0.205 |
| **Swiss language region** | 624 |  |  | 0.36 |
| German-speaking |  | 168 (71%) | 277 (72%) |  |
| French-speaking |  | 62 (26%) | 105 (27%) |  |
| Italian-speaking |  | 7 (3.0%) | 5 (1.3%) |  |
| **Type of workplace** | 624 |  |  | 0.277 |
| University hospital |  | 93 (39%) | 151 (39%) |  |
| Other hospital |  | 88 (37%) | 140 (36%) |  |
| Private practice with <3 colleagues |  | 29 (12%) | 36 (9.3%) |  |
| Private practice with ≥3 colleagues |  | 23 (9.7%) | 42 (11%) |  |
| Other place |  | 4 (1.7%) | 18 (4.7%) |  |
| **Current work quota % (median (IQR))** | 543 | 100 (100, 100) | 100 (80, 100) | <0.001 |
| **Current position** |  |  |  |  |
| General practitioner |  | 49 (21%) | 73 (19%) |  |
| (Dpty) attending physician in GIM |  | 39 (16%) | 54 (14%) |  |
| (Dpty) attending physician in specialty other than GIM |  | 110 (46%) | 212 (55%) |  |
| Resident aiming for certification in GIM |  | 25 (11%) | 29 (7.5%) |  |
| Resident aiming for certification other than GIM |  | 4 (1.7%) | 6 (1.6%) |  |
| Other |  | 49 (21%) | 73 (19%) |  |
| **Currently holding a higher academic degree or position (PhD, lectureship, professorship)** | 536 | 17 (8.5%) | 10 (3.0%) |  |
| **Currently active in research** | 533 | 38 (19%) | 48 (14%) |  |
| **Currently active in medical education** | 533 | 60 (30%) | 78 (23%) |  |
| **Abbreviations: IQR, interquartile range. SD, standard deviation. Dpty, deputy. GIM, general internal medicine. Numbers are presented as n (%), unless indicated otherwise. Missing data were present for currently holding a higher academic career (36 men, 52 women), current activity in research or in medical education (both 36 men, 55 women).** |  |  |  |  |

### Sex differences in career ambitions

Sex-stratified univariate analyses of career aspirations are shown in Table 2. No significant difference existed between the frequency of men and women having had higher career aspirations in the past during medical school (men vs women: 14% vs 12%, p=0.62). However, current career aspirations (after having started their professional career) showed a sex difference, with men reporting higher career aspirations in 17% and women 11% (p=0.03) (Table 2, Supplemental Table 2, Supplemental Table 3). Career ambition scores were higher in men than in women, with 24 (7) (median, IQR) in men and 23 (7) in women (p=0.033). The fraction aiming for an academic position was however similar between the sexes (Table 2). Multivariable logistic regression analyses of the entire population showed that, female sex, the French language -speaking region and working in a group practice were associated with a lower probability of higher career aspirations (Table 3).

**Table 2.** Outcomes.

| **Table 2. Outcomes** |  |  |  |  |
| --- | --- | --- | --- | --- |
|  | **N** | **Men,**<br>**N = 237** | **Women,**<br>**N = 387** | **p value** |
| **Aiming for a Leading Position** | 624 | 40 (17%) | 42 (11%) | 0.031 |
| **Aiming for a Leading Position during medical studies** | 624 | 32 (14%) | 47 (12%) | 0.621 |
| **Career ambition score** | 537 | 24 (7) | 23 (7) | 0.033 |
| **Aiming for an academic career** | 509 | 51 (28%)* | 86 (26%)° | 0.756 |
| **n (%) or mean (standard deviation) is shown. Career ambition score, composite score of 6 questions regarding career ambition with an overall minimum of 6 and maximum 30 points. Missing data occurred in n=87 for Career ambition score. \* number of included men: n=184. ° number of included women: n=325.** |  |  |  |  |

**Table 3.** Factors associated with with higher career aspirations.

| **Table 3: Factors associated with with higher career aspirations** |  |  |  |  |
| --- | --- | --- | --- | --- |
|  |  | **Multivariable logistic regression** |  |  |
| **Characteristic** | **N** | **OR** | **95% CI** | **p-value** |
| **Age** | 625 | 1.07 | 1.03, 1.11 | <0.001 |
| **Swiss language region** | 625 |  |  |  |
| **German-speaking** |  | — | — |  |
| **French-speaking** |  | 0.35 | 0.17, 0.68 | 0.003 |
| **Italian-speaking** |  | 0 |  | >0.99 |
| **Type of work place** | 625 |  |  |  |
| **University hospital** |  | — | — |  |
| **Other place** |  | 0.21 | 0.01, 1.25 | 0.16 |
| **Other hospital** |  | 0.62 | 0.37, 1.03 | 0.066 |
| **Private practice with <3 colleagues** |  | 0 | 0.00, 87.1 | 0.98 |
| **Private practice with ≥3 colleagues** |  | 0.02 | 0.00, 0.10 | <0.001 |
| **Sex** | 625 |  |  |  |
| **Female** |  | — | — |  |
| **Male** |  | 1.74 | 1.06, 2.85 | 0.029 |

OR, odds ratio. CI, confidence interval. The multivariable model contained the variables age, Swiss language region, type of work place and sex. For sex, estimates for nonbinary (n<5) are not shown.

### Sex-stratified analysis of personal and workplace-related factors associated with higher career aspirations

Univariable analyses and multivariable models adjusted for age, Swiss language region and type of workplace are presented in Figures 1-2 and Supplemental Tables 4-5. Female physicians had several personal and workplace-related factors that showed associations with higher career aspirations in multivariable analysis (Figure 1, Supplemental Table 4). Female respondents who were dissatisfied with their work were less likely to have higher career aspirations compared to their counterparts who were satisfied (OR: 0.33,95% CI: 0.09-0.90, p=0.048; Figure 1, Supplemental Table 4). Among women, those with higher career aspirations were more likely to experience negative views on parenthood by their supervisors (OR: 7.48, 95% CI: 1.71-39.0, p=0.01). Finally, in women, higher career aspirations were associated with more frequently reporting gender discrimination by feeling discriminated at work (OR: 2.21, 95% CI: 1.07, 4.53, p=0.031), reporting that the opposite gender receives more responsibilities at work or more recognition for equal achievements (OR: 2.83 [95% CI: 1.37-5.82, p=0.005] and OR: 2.26 [95% CI: 1.11-4.62, p=0.025], respectively). Importantly, having or wanting to have children, having adequate childcare and the distribution of childcare or household duties with one’s partner were not associated with higher career aspirations (Figure 1, Supplemental Table 4). In addition, answers to questions regarding workplace inclusiveness were also not associated with aiming for a leading position. A subgroup analysis of physician-mothers yielded overall similar results (Supplemental Table 6).

**Figure 1:**
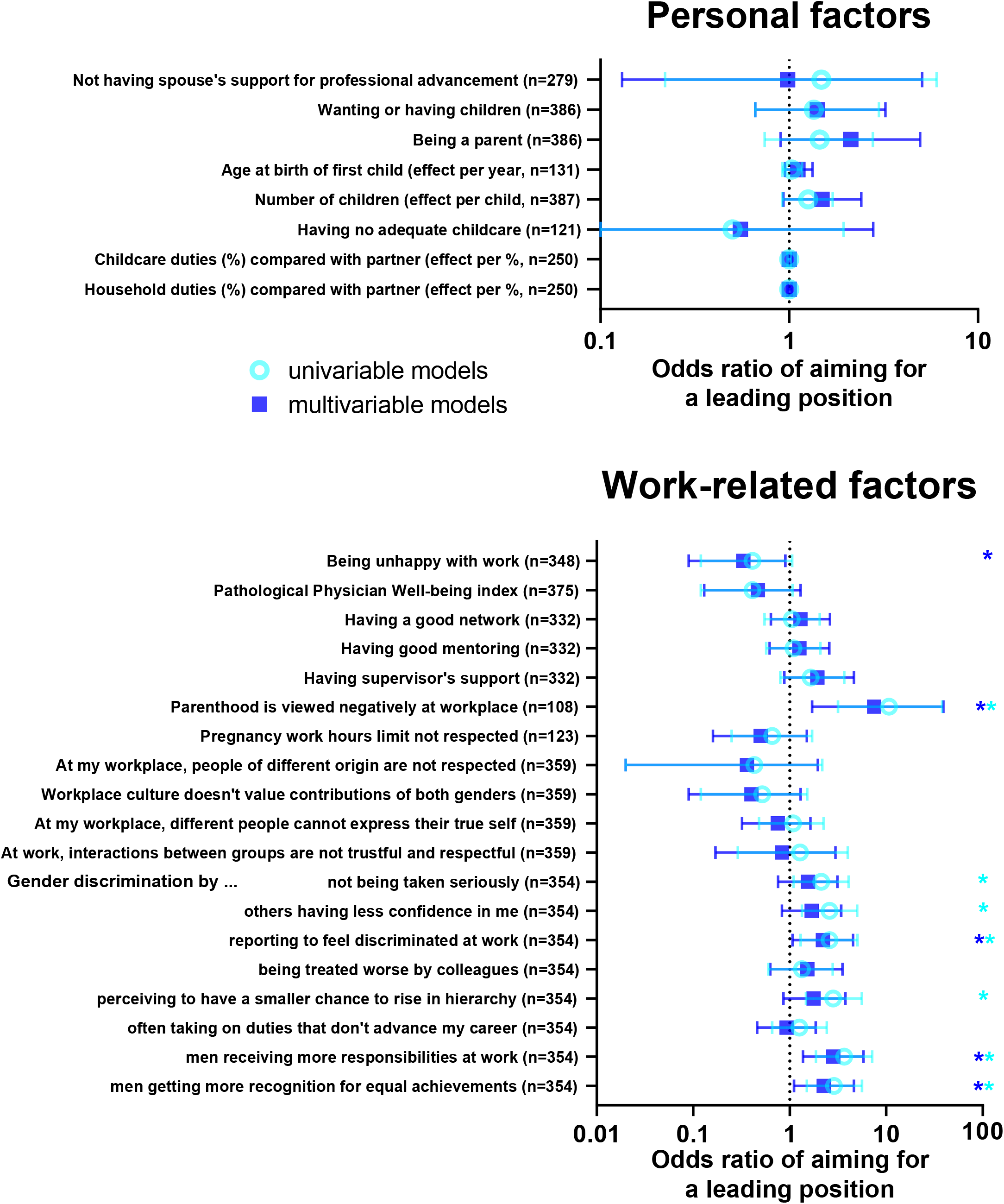
Higher career aspirations among female physicians. Forest plot of the odds ratios of personal (top panel) and work-related factors (lower panel) for higher career aspirations among female physicians. Each row depicts an univariable model (light blue) and a multivariable logistic regression model (dark blue) adjusted for age, Swiss language region and type of workplace. Stars indicate significant associations, and error bars indicate 95% confidence intervals.

**Figure 2:**
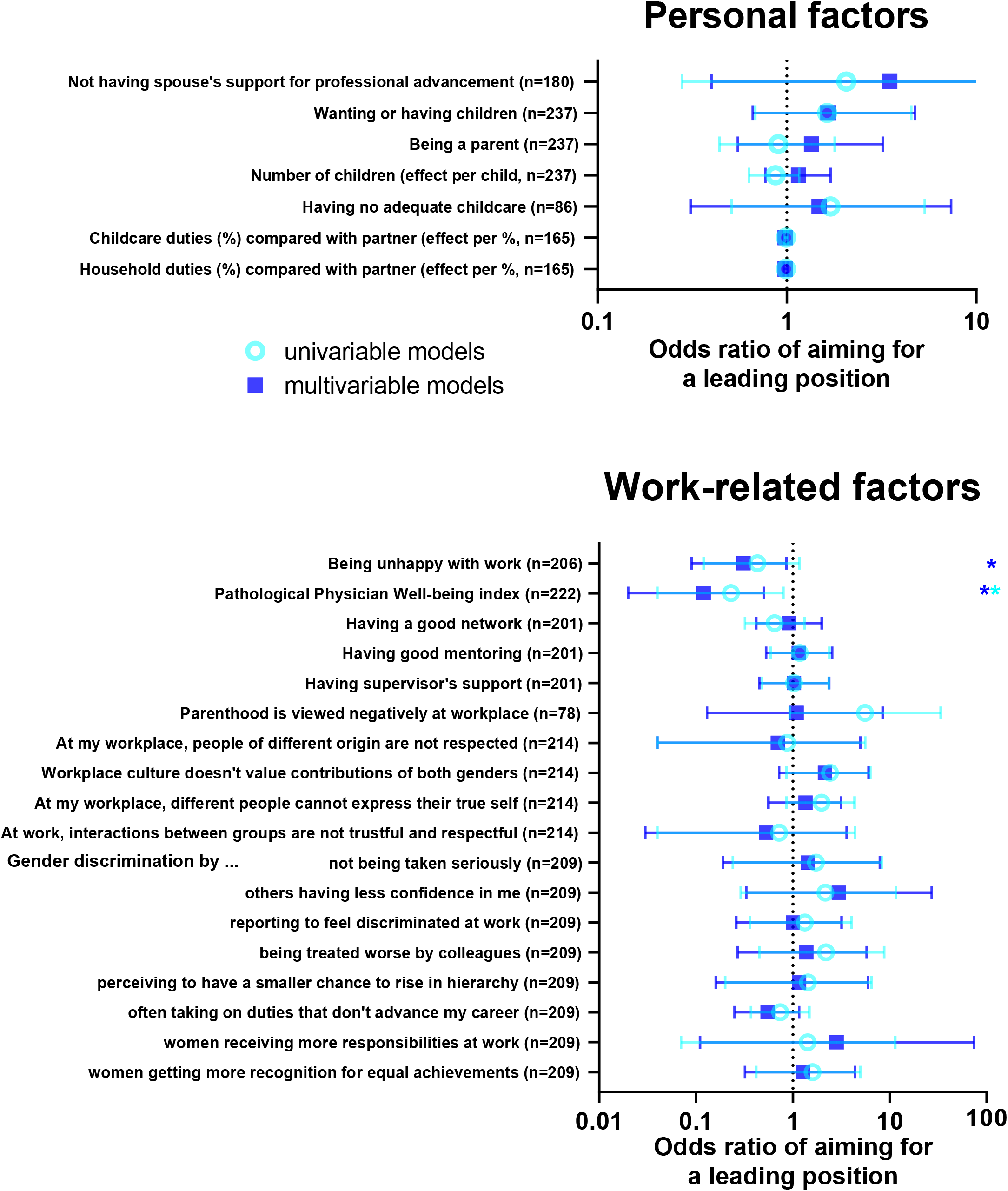
Higher career aspirations among male physicians. Forest plot of the odds ratios of personal (top panel) and work-related factors (lower panel) for higher career aspirations among male physicians. Each row depicts an univariable model (light blue) and a multivariable logistic regression model (dark blue) adjusted for age, Swiss language region and type of workplace. Stars indicate significant associations, and error bars indicate 95% confidence intervals.

In male physicians, a different summary of personal factors but not workplace-related factors were associated with higher career aspirations (Figure 4, Supplemental Table 5): Similar to female participants, male physicians who were dissatisfied with work also showed a lower probability of higher career aspirations (OR 0.31, 95% CI: 0.09-0.86, p=0.04). However, male physicians who had a pathological PWBI were less likely to have higher career aspirations, with an OR 0.12 (95% CI: 0.02-0.50, p=0.012). In the male group, responses covering gender discrimination or workplace inclusiveness were not associated with higher career aspirations (Figure 4, Supplemental Table 5). A subgroup analysis including physician-as fathers identified no associations with higher career aspirations in adjusted analysis (Supplemental Table 7).

### Sex-stratified analyses of factors associated with aiming for an academic career and career ambition scores

In women, aiming for an academic career was associated with not complying with the maximum legally allowed weekly working hours during pregnancy (OR 3.65 (95% CI: 1.09-14.8, p=0.046 p > 0.05; Supplemental Table 8). Women aiming for an academic career had a high likelihood of reporting gender discrimination in several instances, such as not being taken seriously, receiving less confidence, being treated worse by colleagues, and often taking on duties that were not advancing their career (Supplemental Table 8). Among women, being a parent was associated with a lower probability of aiming for an academic career only in univariable but not in adjusted analysis (Supplemental Table 8). Among men, working at an institution that reportedly does not value both genders’ contributions was associated with aiming for an academic career (OR 3.57 (1.17-11.5, p=0.027), Supplemental Table 9).

Regarding career ambition, feeling unhappy because of work was associated with lower partial CAS-R scores among women, whereas a higher rating in the partial CAS-R scores was associated with frequent incidence of perceived gender-related discrimination (Supplemental Table 10). Career ambition scores were higher in male respondents when factors were present such as having support by superiors, having a good network and good mentoring (Supplemental Table 11).

### Sensitivity analysis

A sex-stratified result of an analysis excluding general practitioners were overall comparable to the main analysis (Supplemental Tables 12-13). This analysis strengthens the robustness of the factors associated with higher career aspirations in the present dataset.

## Discussion

In this study, we aimed to describe career aspirations of physicians in GIM in Switzerland and to verify which personal and work-related factors are associated with higher career aspirations. Overall, female and male physicians had similar career aspirations during medical studies, but career aspirations appeared to decrease over time in female physicians. Among physicians of both sexes, dissatisfaction with work is associated with a lower probability of aiming for a leading position. Finally, gender discrimination was identified as an important workplace-related factor with a potential for career obstruction when female physicians aimed to pursue a career path towards hospital leadership or towards an academic career.

Our findings of an apparent decline in female physicians’ career ambitions over the time strengthen the existing evidence from other countries. A German study demonstrated no evidence of a gap in career ambition between women and men during early medical studies, but a career ambition gap occurring both among advanced medical students and physicians.(32) Further, gender discrimination related to pregnancy and parenthood was reported as the most common cause of discrimination among female physicians in Switzerland.(24) Furthermore, in female non-physician academics in Switzerland, even the potential future first-time motherhood was associated with negative workplace experiences.(33) In contrast, women’s ambitions remained high and comparable to men’s over the time if the work environment was supportive for several professions.(15) Our findings align with recent U.S. data suggesting that female faculty is more likely to report to be pushed from their academic positions than male.(34) Although no direct causality can be established in an observational study design, the present findings and the literature support the notion that gender discrimination remains an important issue associated with the decreased career aspirations in women, and also significantly disadvantaging women in work assignments or promotions. As consequence, this leads to a reduced fraction of women attaining leadership positions.(4,24) Our findings support the need and implications for a tailored strategies in national and international interventions directed towards optimization of the physician workplace environment, thus allowing to fully mobilize the human capital and retain potential female leadership.(35)

Strengths of this study include the comprehensive characterization of both personal and work-place related factors in association with higher career ambitions. Furthermore, the study included a large sample of 624 participants, constituting roughly 1.5% of all practicing physicians in Switzerland. Finally, the present study allows to disentangle sex differences, as the questionnaires were answered by respondents of a relatively balanced sex ratio. Further, the observed differences in factors associated with career aspirations among female and male physicians – a focus on overcoming negative views on motherhood and preventing gender-related discrimination – could likely give a strong incentive for guiding specific interventions to promote an organizational change in hospitals in Switzerland.(36)

The present study also has some limitations. First, the survey response rate cannot be determined, as the number of viewers of the survey advertisements is unknown and website views were not measured. However, the PWBI prevalence among the present data were similar to previous Swiss studies with response rate of 40% (29,30). This indicates that, for instance, physicians who are unwell were not overrepresented among the present sample. Next, different career stages cannot directly be longitudinally compared due to the cross-sectional design of the study. In addition, causality could not be detected based on the reported associations alone, and as is the case in all observational studies. Finally, this survey was limited to Switzerland, and therefore the results may not necessarily be generalizable to countries with different education and health care systems.

Based on the outcomes of this study, several further steps can be formulated that could be considered and evaluated. First, work environment policies would need to be adapted towards implementation of educational activities to understand the roots of disadvantage of losing the contribution of incentives for female leadership in health care sector. Irrespectively of any government’s involvement, the current actions could be initiated even during medical education trying to build career ladder models, including leadership courses, for the future generation of physicians which is dedicated to improve gender equality issues towards a progress for better health and its associated services. Current leaders in medicine might potentially also need some educational activity for a deeper understanding of existing problems and to gain potential tools that could help to minimize gender bias during recruitment process or in the following advancement of career development. Finally, a still widely debated topic of strategic recruitment of only female leadership might contribute to certain environment and certain time, although this must be considered with caution by not excluding the professional and soft skill competence needed for future tasks.

## Conclusion

Female and male physicians equally aim for leading positions at their work at the beginning of their career in medicine, but the different types and intensity of challenges that women and men face during their career progression could cause or amplify the underrepresentation of women among leadership for medical health care providers. Interventions to diminish discrimination related to parenthood or gender among GIM physicians could contribute to increased inclusivity and preserve career aspirations, and such interventions should be carefully designed and evaluated in prospective studies.

## Acknowledgements

The authors are thankful to the respondents, and to the support by Swiss Society of Internal Medicine, Swiss Young General Practitioners Association (JHaS), Primary Hospital Care and the hospitals involved with distribution of the survey.

## Data availability

All data produced in the present study are available upon reasonable request to the authors.

